# Use of Lecanemab and Donanemab in the Canadian Healthcare System: Evidence, Challenges, and Areas for Future Research

**DOI:** 10.1101/2024.11.18.24317292

**Authors:** Eric E. Smith, Natalie Phillips, Howard H. Feldman, Michael Borrie, Aravind Ganesh, Alexandre Henri-Bhargava, Philippe Desmarais, Andrew Frank, AmanPreet Badhwar, Laura Barlow, Robert Bartha, Sarah Best, Jennifer Bethell, Jaspreet Bhangu, Sandra E. Black, Christian Bocti, Susan E. Bronskill, Amer M. Burhan, Frederic Calon, Richard Camicioli, Barry Campbell, D. Louis Collins, Mahsa Dadar, Mari L. DeMarco, Simon Ducharme, Simon Duchesne, Gillian Einstein, John D. Fisk, Jodie R. Gawryluk, Linda Grossman, Zahinoor Ismail, Inbal Itzhak, Manish Joshi, Arthur Harrison, Edeltraut Kroger, Sanjeev Kumar, Robert Laforce, Krista L. Lanctot, Meghan Lau, Linda Lee, Mario Masellis, Fadi Massoud, Sara B. Mitchell, Manuel Montero-Odasso, Karen Myers, Haakon B. Nygaard, Stephen H. Pasternak, Jody Peters, M. Natasha Rajah, Julie M. Robillard, Ken Rockwood, Pedro Rosa-Neto, Dallas P. Seitz, Jean-Paul Soucy, Shanna C. Trenaman, Cheryl L. Wellington, Aicha Zadem, Howard Chertkow, the Canadian Consortium on Neurodegeneration in Aging Investigators

## Abstract

Lecanemab and donanemab are monoclonal antibody therapies that remove amyloid-beta from the brain. They are the first therapies that alter a fundamental mechanism, amyloid-beta deposition, in Alzheimer disease (AD). To inform Canadian decisions on approval and use of these drugs, the Canadian Consortium on Neurodegeneration in Aging commissioned Work Groups to review evidence on the efficacy, and safety of these new therapies, as well as their projected impacts on Canadian dementia systems of care. We included persons with lived experience with Alzheimer disease in the discussion about the benefits and harms. Our review of the trial publications found strong support for statistically significant group differences, but also recognized that there are mixed views on the clinical relevance of the observed differences and the value of therapy for individual patients. The drugs are intended for persons with early AD, at a stage of mild cognitive impairment or mild dementia. If patients are treated, then confirmation of AD by positron emission tomography or cerebrospinal fluid analysis and monitoring for risk of amyloid-related imaging abnormalities was recommended, as done in the clinical trials, although it would strain Canadian resource capacity. More data are needed to determine the size of the potentially eligible treatment population in Canada.

## INTRODUCTION

Alzheimer disease (AD) is a progressive, neurodegenerative disease that causes cognitive decline and, ultimately, death(1). AD is marked by accumulation of plaques, composed of amyloid-beta, and neurofibrillary tangles, composed of tau, in the brain. The only Health Canada approved drugs to treat AD are the cholinesterase inhibitors and memantine, which have been shown to enhance cognition but do not influence accumulation of amyloid-beta(2). In 2023, phase 3 randomized controlled trials (RCTs) showed that lecanemab(3) and donanemab(4), monoclonal antibodies targeted against amyloid-beta, reduced the rate of cognitive and functional decline in persons with AD compared with placebo. Both drugs are now approved by the United States (US) Food and Drug Administration (FDA) and reimbursed by US Medicare, conditional on reporting information to an approved registry. Previously, and controversially, a drug from the same class, aducanumab, was conditionally approved by the US FDA for removal of amyloid-beta from the brain despite mixed results from two RCTs(5); however, the drug was rarely prescribed and it is no longer produced by the manufacturer.

The clinical benefits, harms, and cost effectiveness of these drugs have been controversial. In contrast to the US FDA decision, the European Medicines Agency initially declined to approve lecanemab for use in Europe, citing significant harms as well as meager benefits, but then reversed its decision on November 14, 2024, approving lecanemab for treatment of patients with zero or one copy, but not two copies, of the *APOE* ε4 allele. In the UK, the Medicines and Healthcare products Regulatory Agency (MHRA) granted approval for marketing of lecanemab and donanemab for patients with zero or one copy of the *APOE* ε4 allele. but the National Institute for Health and Care Excellence has issued draft guidance recommending against coverage by the National Health Service. In contrast, at the time of writing lecanemab has been approved in China, Israel, Japan, South Korea, and the United Arab Emirates regardless of *APOE* genotype; and donanemab has been approved in Japan. In contrast, the Australian Therapeutics Goods Administration declined to approve lecanemab.

Lecanemab and donanemab are currently under review by Health Canada, which issues approval for marketing drugs in a similar fashion as the US FDA, and the Canadian Drug Agency, which will issue a report on the drugs, including their cost effectiveness, that will be used by provincial formularies to decide whether to reimburse the costs of the drugs in each province.

Previous reports have suggested that the Canadian healthcare system is ill prepared for disease-modifying drugs for AD, with significant barriers to accessing diagnostic testing and speciality care(6,7). While these reports were sponsored by pharmaceutical companies, which could raise concerns over potential conflict of interest, similar concerns about health system readiness have been shared by Canadian editorialists from the academic community(8-10). Additionally, the clinical value of the drug effects, balanced against the risks, has engendered much debate(11).

The Canadian Consortium on Neurodegeneration in Aging (CCNA) is Canada’s nationally funded dementia research network(12). To inform decisions on the utility and feasibility of lecanemab and donanemab, the CCNA commissioned a contemporary review of the effectiveness of these therapies, how they could be applied in the clinic, challenges with their potential use in the Canadian healthcare system, and a future research agenda. The primary intent of this review is to convey contemporary information on the efficacy and clinical use of these drugs to clinicians, health system administrators, regulators, and policy makers in Canada and elsewhere.

## METHODS

The CCNA Research Executive Committee, the main decision-making body of the CCNA, commissioned a Steering Committee for this initiative on June 7, 2024. Nine Work Groups were created by soliciting volunteers from among the 320 co-investigators of the CCNA. Additionally, to obtain the perspectives of patients and caregivers, members of the Engagement of People with Lived Experience(13) were recruited to participate in a focus group session on the benefits and risks of treatment.. Members of the writing committee were required to disclose all relevant financial and professional conflicts of interest, following policies of the International Committee of Medical Journal Editors. The Chair of the Steering Committee (Smith) was required to be free of conflicts during the work of the initiative; otherwise, potential conflicts were not considered to be disqualifying but had to be disclosed when discussing any issue that might overlap with their personal or professional interests.

Work Group members reviewed peer-reviewed publications, supplemental files, and protocols of the CLARITY-AD (lecanemab)(3) and TRAILBLAZER-ALZ 2 (donanemab)(4) trials. Peer-reviewed data published prior to November 7, 2024, were prioritized. When needed, Plot Digitizer was used to extract numeric data from figures(14).

## CLARITY-AD AND TRAILBLAZER-ALZ 2: TRIAL DESIGNS

### Clinical trials of lecanemab and donanemab

The CLARITY-AD trial(3) tested the hypothesis that lecanemab, compared with placebo, would reduce the rate of decline on the Clinical Dementia Rating Sum of Boxes (CDR-SB) over 18 months(15). The CDR-SB is derived from a structured interview with both the participant and an informant, rating performance in six domains: memory, orientation, judgment and problem solving, community activities, home and hobbies, and personal care(15). The design of CLARITY-AD is shown in Figure 1A. Lecanemab is a monoclonal antibody targeted against amyloid-beta protofibrils(16). It was infused intravenously every 2 weeks at a dose of 10 mg/kg. The trial duration was 18 months, with an open-label extension period thereafter. Periodic magnetic resonance imaging (MRI) scans were required to monitor for amyloid related imaging abnormalities (ARIA).

**Figure 1.**
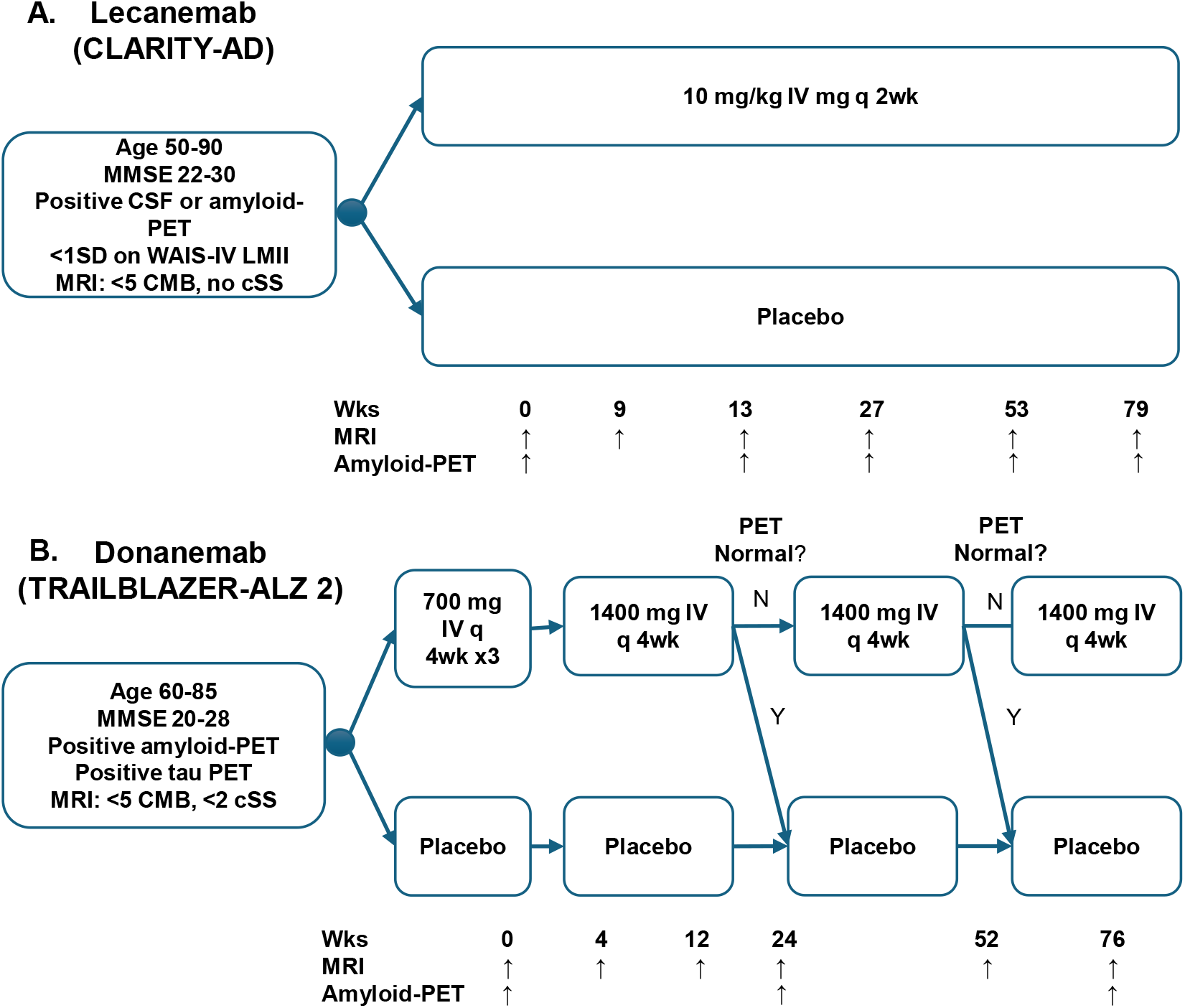
Design of the CLARITY-AD (Lecanemab) and TRAILBLAZER-ALZ2 (Donanemab) Clinical Trials. In TRAILBLAZER-ALZ 2, participants on donanemab crossed over to the placebo arm at 24 weeks or 52 weeks if the amyloid-PET was <11 Centiloids on any single PET scan or ≥11 but <25 Centiloids on 2 consecutive PET scans. CMB, cerebral microbleed; cSS, cortical superficial siderosis; CSF, cerebrospinal fluid; MMSE, Folstein Mini-Mental State Examination; MRI, magnetic resonance imaging; PET, positron emission tomography; WAIS-IV LMII, Wechsler Adult Intelligence Scale IV Logical Memory II test; Wks, weeks.

The TRAILBLAZER-ALZ 2 trial(4) tested the hypothesis that donanemab, compared with placebo, would reduce the rate of decline on the integrated Alzheimer’s Disease Rating Scale (iADRS) over 18 months. The iADRS is a combination of scores from two widely used measures in AD trials: the Alzheimer’s Disease Assessment Scale Cognitive Subscale (ADAS-Cog; a battery of neuropsychological tests) and the Alzheimer’s Disease Cooperative Study – instrumental Activities of Daily Living scale (ADCS-iADL)(17). Thus, the primary outcome measure differed from the CLARITY-AD trial of lecanemab; however, the change in CDR-SB was reported as a secondary outcome measure, allowing comparison on the same outcome across the two trials. The study design is shown in Figure 1B. Donanemab is a monoclonal antibody targeted against plaque amyloid(18). It was infused intravenously every 4 weeks, beginning at 700 mg IV each month for 3 months and then increased to the target dose of 1400 mg IV every month. The schedule of MRIs is shown in Figure 1B. Randomization was stratified by level of tau, measured by positron emission tomography (PET): “low-medium tau” (meaning that tau was only mildly or moderately elevated above normal) and “high tau” (more than moderately elevated). According to the trial protocol, there were two primary outcomes: change in iADRS in the low-medium tau group, and change in iADRS in the combined tau group, pooling the low-medium and high groups. A unique feature of TRIALBLAZER-ALZ2 was that donanemab infusions were stopped if follow-up amyloid-PET signal normalized, which was achieved in 29.7% of participants at the 6-month follow-up and 76.4% of participants at the 12-month follow-up.

In the remainder of this document, we will collectively refer to lecanemab and donanemab as anti-amyloid-beta monoclonal antibodies (anti-Aβ mAbs).

## BENEFITS AND HARMS OF TREATMENT

### Clinical meaning of the benefits

No aspect of the trials has engendered more controversy than the clinical value of the drug effects on participant well-being and quality of life. In the clinical trials, across all participants the relative change in cognitive and functional decline was proportionally much less than the relative reduction in amyloid-PET signal(3,4). The reasons for this discrepancy are unknown, but could include lack of effect on soluble precursors to amyloid (e.g., oligomeric Aβ) that are not measured by existing biomarkers, feed-forward neurodegenerative loops that were triggered by amyloid-beta but can no longer be interrupted by its removal, unexpectedly high contributions of comorbidities that are independent of amyloid-beta (including vascular, Lewy body, and other pathologies), or subtle adverse cognitive effects of the drugs. Research is urgently needed to determine the cause of this discrepancy, with important implications for the value that can ultimately be expected of this drug class.

The effects of these medications on the primary and important secondary study outcomes are shown in Table 1(3,4,19). In the primary analysis of the CLARITY-AD trial of lecanemab, the adjusted least-squares mean change from baseline at 18 months (more positive scores are worse) was 1.21 with lecanemab and 1.66 with placebo (difference −0.45; 95% CI −0.67 to −0.23, p<0.001). In TRAILBLAZER-ALZ 2 outcomes were reported separately for the low/medium tau population and the combined population, which included high tau as well as low/medium tau(4). In the primary analysis, the adjusted least-squares mean change from baseline at 18 months (more negative is worse) in donanemab compared with placebo in the low/medium tau population was -6.02 versus -9.27 (difference +3.25, 95% CI +1.88 to +4.62, p<0.001; relative difference 35.1%) and in the combined tau population was -10.19 versus -13.11 (difference +2.92, 95% CI 19.9-50.2%, 95% CI +1.51 to +4.33, p<0.001; relative difference 22.3%). In both trials, the models included terms for the interaction of treatment by visit, which tests the difference in slopes between treatment and placebo, but these model coefficients were not reported.

**Table 1.** Primary Outcome and Selected Secondary Outcomes of CLARITY-AD and TRAILBLAZER-ALZ 2. *p<0.05 **p<0.01 ***p<0.001. Values are adjusted least-squares mean change from baseline at 18 months and their differences between treatment and placebo, except for time to global CDR change which is give as the percent with an increase to a higher global CDR at 18 months, the risk difference, and hazard ratio. Values are scale points, except for amyloid removal which is in centiloids. Numbers in brackets are 95% confidence limits; however, 95% confidence limits were not reported for all analyses. Time to global CDR change is based on a change from baseline to a higher global CDR (global CDR in trial participants could have values of 0.5, 1.0, 2.0 or 3.0; corresponding to questionable, mild, moderate, or severe dementia). ADAS-Cog, Alzheimer's Disease Assessment Scale-Cognitive Subscale; ADCS MCI-ADL, Alzheimer’s Disease Cooperative Study-Activities of Daily Living Scale for Mild Cognitive Impairment; ADCS-IADL Alzheimer’s Disease Cooperative Study-Instrumental Activities of Daily Living; CDR-SB, Clinical Dementia Rating Sum of Boxes; EQ-5D-5L, EuroQol 5D-5L; iADRS, integrated Alzheimer’s Disease Rating Scale; MMSE, Folstein MiniMental Status Exam; QOL-AD, Quality of Life-Alzheimer Disease scale; ZBI, Zarit Burden Interview.

| Outcome | CLARITY-AD<br>(Lecanemab) | Mean | 95% CI | TRAILBLAZER-ALZ 2<br>(Donanemab) | Mean | 95% CI |
| --- | --- | --- | --- | --- | --- | --- |
| Primary | **CDR-SB** |  |  | **iADRS** |  |  |
|  |  |  |  | **Low-medium tau** |  |  |
|  | Lecanemab | +1.21 | -- | Donanemab | -6.02 | -7.01 to -5.03 |
|  | Placebo | +1.66 | -- | Placebo | -9.27 | -10.23 to -8.31 |
|  | Difference | -0.45*** | (-0.67 to -0.23) | Difference | +3.25 | 1.88- to .62*** |
|  | Slowing | 27.1% | -- | Slowing | 35.1% | 19.9-50.2% |
|  |  |  |  | **Combined** |  |  |
|  |  |  |  | Donanemab | -10.2 | -11.2 to -9.32 |
|  |  |  |  | Placebo | -13.1 | -14.1 to -12.13 |
|  |  |  |  | Difference | +2.92 | 1.51-4.33*** |
|  |  |  |  | Slowing | 22.3% | 11.49-33.2% |
| **CDR-SB** | See above |  |  | **Low-medium tau** |  |  |
|  |  |  |  | Donanemab | +1.20 | 1.00 to 1.41 |
|  |  |  |  | Placebo | +1.88 | 1.68 to 2.08 |
|  |  |  |  | Difference | -0.67*** | -0.95 to -0.40 |
|  |  |  |  | Slowing | 36.0% | 20.8-50.2% |
|  |  |  |  | **Combined** |  |  |
|  |  |  |  | Donanemab | +1.72 | 1.53 to 1.91 |
|  |  |  |  | Placebo | +2.42 | 2.24 to 2.60 |
|  |  |  |  | Difference | -0.70 | -0.95 to -0.45 |
|  |  |  |  | Slowing | 28.9% | 18.3-39.5% |
| **Time to CDR Increase** |  |  |  | **Low-medium** |  |  |
|  | Lecanemab | 31.9% | -- | Donanemab | 30.3% | -- |
|  | Placebo | 23.4% | -- | Placebo | 19.9% | -- |
|  | Risk diff | 8.5% | -- | Risk difference | 10.4% | -- |
|  | Hazard ratio | 0.69 | -- | Hazard ratio | 0.61 | 0.47-0.80 |
|  |  |  |  | **Low-medium** |  |  |
|  |  |  |  | Donanemab | 36.4% | -- |
|  |  |  |  | Placebo | 25.6% | -- |
|  |  |  |  | Risk difference | 10.8% | -- |
|  |  |  |  | HR | 0.63 | 0.51 to 0.77 |
| **Activities of Living** | **ADCS-MCI-ADL** |  |  | **ADCS-IADL** |  |  |
|  |  |  |  | **Low-medium tau** |  |  |
|  | Lecanemab | -3.5 | -- | Donanemab | -2.76 | -3.42 to -2.10 |
|  | Placebo | -5.5 | -- | Placebo | -4.59 | -5.23 to -3.95 |
|  | Difference | +2.0*** | (1.2 to -2.8) | Difference | +1.83*** | 0.91 to 2.75 |
|  |  |  |  | Slowing | 39.9% | 19.2-60.6% |
|  |  |  |  | **Combined** |  |  |
|  |  |  |  | Donanemab | -4.42 | -5.05 to -3.80 |
|  |  |  |  | Placebo | -6.13 | -6.72 to -5.53 |
|  |  |  |  | Difference | +1.70*** | 0.8 to 2.6 |
|  |  |  |  | Slowing | 27.8% | 13.4-42.1% |
| **MMSE** | Not reported |  |  | **Low-medium tau** |  |  |
|  |  |  |  | Donanemab | -1.61 | -1.89 to -1.33 |
|  |  |  |  | Placebo | -2.09 | -2.36 to -1.81 |
|  |  |  |  | Difference | +0.48* | 0.09 to 0.87 |
|  |  |  |  | Slowing | 22.9% | 4.0-41.8% |
|  |  |  |  | **Combined** |  |  |
|  |  |  |  | Donanemab | -2.47 | -2.73 to -2.20 |
|  |  |  |  | Placebo | -2.94 | -3.20 to -2.69 |
|  |  |  |  | Difference | +0.47* | 0.10 to 0.84 |
|  |  |  |  | Slowing | 16.1% | 3.5-28.7% |
| **ADAS-Cog** | Lecanemab | +4.14 | -- | **Low-medium tau** |  |  |
|  | Placebo | +5.58 | -- | Donanemab | +3.17 | 2.64 to 3.69 |
|  | Difference | -1.44*** | -2.27 to -0.61 | Placebo | +4.69 | 4.18 to 5.20 |
|  |  |  |  | Difference | -1.52*** | -2.25 to -0.79 |
|  |  |  |  | Slowing | 32.4% | 16.6-48.4% |
|  |  |  |  | **Combined** |  |  |
|  |  |  |  | Donanemab | +5.46 | 4.91 to 6.01 |
|  |  |  |  | Placebo | +6.79 | 6.26 to 7.32 |
|  |  |  |  | Difference | -1.33*** | -2.09 to -0.57 |
|  |  |  |  | Slowing | 19.5% | 8.2-30.8% |
| **Amyloid removal** | Lecanemab | -55.5 | -- | **Low-medium tau** |  |  |
|  | Placebo | +3.64 | -- | Donanemab | -88.0 | -- |
|  | Difference | -59.1*** | -62.6 to -55.6 | Placebo | +0.2% | -- |
|  |  |  |  | Difference | -88.2*** | -- |
|  |  |  |  | **Combined** |  |  |
|  |  |  |  | Donanemab | -87.0 | -- |
|  |  |  |  | Placebo | -0.7 | -- |
|  |  |  |  | Difference | -86.3*** | -- |
| **Quality of Life** | EQ-5D-5L |  |  |  |  |  |
|  | Lecanemab | -2.12 | -2.61 to -1.58 |  |  |  |
|  | Placebo | -4.13 | -4.63 to -3.60 |  |  |  |
|  | Difference | +2.0** | -- |  |  |  |
|  | Slowing | 49% | -- |  |  |  |
|  | QOL-AD Participant |  |  |  |  |  |
|  | Lecanemab | -0.52 | -0.70 to -0.37 |  |  |  |
|  | Placebo | -1.19 | -1.35 to -1.02 |  |  |  |
|  | Difference | +0.66** | -- |  |  |  |
|  | Slowing | 56% | -- |  |  |  |
|  | QOL-AD Partner |  |  |  |  |  |
|  | Lecanemab | -1.82 | -1.99 to -1.63 |  |  |  |
|  | Placebo | -2.34 | -2.52 to -2.18 |  |  |  |
|  | Difference | +0.54* | -- |  |  |  |
|  | Slowing | 23% | -- |  |  |  |
| **Caregiver ZBI** | Lecanemab | 3.58 | 3.21 to 3.97 |  |  |  |
|  | Placebo | 5.79 | 5.41 to 6.13 |  |  |  |
|  | Difference | -2.2*** | -- |  |  |  |

Some expert opinion has been skeptical of the clinical value of the effects. The European Medicine Agency, to justify their initial decision to decline approval of lecanemab (which was subsequently reversed), stated that the “difference between the two groups was small”. Experts have also noted that treatment may not be desirable for patients who dislike interacting with the health care system or do not live near a major academic medical center(8,20). But for individuals who are eager to slow the disease, even without large benefits on quality of life, the benefits compared with the risks may be acceptable. Research on patient, caregiver, and public opinion has so far been limited.

Experts have noted that the differences between treatment and placebo groups in the trials were less than previously derived minimal clinically important differences (MCIDs) for the primary outcomes. However, MCIDs are, by design, estimates of thresholds for meaningful change within a single individual, and should not be used to judge the clinical value of group average differences(21). To judge the trial outcomes on their MCIDs, one would need to identify the proportion of individuals in each group that did or did not exhibit a meaningful change, which so far has not been reported. There may be a minority of participants who would be considered individual responders, with larger, more clearly meaningful clinical benefits. Another reported method for expressing the meaning of group differences is in “time saved”(22). Analyses of CLARITY-AD and TRAILBLAZER-ALZ 2 suggest that the time saved on the iADRS was 5.3 months over 18 months of treatment, meaning that at the end of 18 months the treated group on average had a CDR change that was the same as what the placebo group had experienced 5.3 months earlier(22).

Future research on clinical effects should include more comprehensive assessment of effects across different neuropsychological domains, participant-reported quality of life, caregiver burden, caregiver health and quality of life, and how these relate to changes in cognition and CDR-SB(23-25). Improved quality of life for both participant and study partner were seen in the CLARITY-AD trial of lecanemab(19). Fuller reporting of trial primary and secondary outcomes, including the confidence limits around change over time in CLARITY-AD and interactions between treatment and time for both trials, is encouraged.

A disease-modifying treatment that alters the slope of decline will continue to accrue benefit the longer it is applied, assuming the slopes of decline remain linear over time. However, to date the only published data are from the first 18 months of treatment in the randomized trials, and the slopes of decline have not been compared statistically to quantify their divergence. It will be critically important to collect data from open label extensions of these trials, and from routine practice, to evaluate the slope of decline with longer term treatment. It is also important to collect data on rate of decline after treatment is stopped, including when treatment is stopped because the amyloid-beta is cleared as was done in TRAILBLAZER-ALZ 2.

### Clinical effect: perspective of persons with lived experience

Recent developments in anti-Aβ mAb therapies were discussed with people with lived experience of dementia through the CCNA Engagement of People with Lived Experience of Dementia (EPLED) program(13). Contributors included a person living with dementia, and four current and former caregivers and care partners, including one who took part in a trial of another disease modifying therapy. The contributors were offered pre-readings(8,20) and took part in a 90-minute discussion about the therapies. Themes that emerged included a desire for patient and care partner choice, but also concerns regarding quality of life, access to a variety of care options, and lack of resources.

Decisions around engaging with new anti-Aβ mAb therapies were seen as personal and to depend on, among many factors, life-stage, disease stage, caregiver support, availability and accessibility of services and, potentially, ability to pay for treatment (e.g., private insurance). These factors must be considered in individual decision-making, but some must also be considered as potential indicators of structural and social determinants of health that might result in inequitable access to care.

Any clinical benefits derived from disease modifying therapy in early/mild stage dementia must be considered against the potential to also prolong later stage dementia, when the symptoms are severe and caregiver burden is high. Key considerations for developing and implementing new therapies were seen to include determining their long-term impacts on an individual’s symptoms and quality of life, as well as their caregiver health and wellbeing, to inform when and how to also discontinue treatment.

Participants expressed concern that dementia care is underfunded and without substantial increased investment across multiple areas of the health and social care system (e.g., home care, long-term care, palliative care and caregiver support), focused investment in the infrastructure that would be required to support treatment with anti-Aβ mAbs was not seen as the best use of limited resources.

Many individuals and families already face challenges in obtaining a timely dementia diagnosis. Given that treatment is contingent on early diagnosis (and ongoing monitoring), potential for existing gaps and disparities in health service use(26,27) to extend to effective treatment with anti-Aβ mAbs must be addressed.

Finally, there were calls to invest in research on more effective therapies, even though investment so far has been mostly disappointing, with many failures. However, these failures were also seen to highlight the imperative to not only advance therapies, but also to develop and implement effective approaches to primary prevention as well as non-pharmacological services and supports for persons living with dementia and their caregivers.

### Amyloid related imaging abnormalities and other adverse effects

Amyloid-related imaging abnormalities (ARIA) are the most important potential adverse effects of treatment with anti-Aβ mAbs. In addition to ARIA, headache (possibly because of ARIA) and infusion reactions (discussed in more detail in the section on Organizing Care) were more common with treatment than placebo.

ARIA refers to MRI evidence of vasogenic edema or bleeding that can occur in response to anti-Aβ mAbs. A consensus group convened by the Alzheimer’s Association defined ARIA with edema (ARIA-E) as “MRI alterations thought to represent edema in the gray and white matter, and effusion or extravasated fluid in the sulcal space (28). ARIA with hemorrhage (ARIA-H) was defined as MRI findings thought to represent hemosiderin deposits, including microbleeds, macrohemorrhages, and superficial siderosis(28).

The incidence of ARIA-E and ARIA-H in the CLARITY-AD and TRAILBLAZER-ALZ 2 trials is shown in Table 2. The rate of ARIA-E and ARIA-H was higher in donanemab-treated participants than lecanemab-treated participants; however, this difference should be viewed cautiously because the two drugs were not compared head-to-head in the same trial. When ARIA occurred, it was usually early in the treatment course: in TRAILBLAZER-ALZ 2, 58% of first instances of ARIA-E occurred within the first three doses(4). ARIA also occurred in the placebo groups: ARIA-E was about 1/10^th^ as common, and ARIA-H was about half as common in placebo compared with treatment.

**Table 2.** Rates of Amyloid-Related Imaging Abnormalities (ARIA)

| **ARIA type** | **CLARITY-AD** |  | **TRAILBLAZER-ALZ 2** |  |
| --- | --- | --- | --- | --- |
|  | **Lecanemab** | **Placebo** | **Donanemab** | **Placebo** |
| Any ARIA (-E or -H) | 21.5% | 9.5% | 36.8% | 14.9% |
| ARIA-E (radiological) | 12.6% | 1.7% | 24.0% | 1.9% |
| ARIA-E (symptomatic) | 2.8% | 0.0% | 6.1% | 0.1% |
| ARIA-H (radiological, all types) | 17.3% | 9.0% | 31.4% | 13.6% |
| New microbleeds | 14.0% | 7.6% | 26.8% | 12.5% |
| New superficial siderosis | 5.6% | 2.3% | 15.7% | 3.0% |
| New macrohemorrhages | 0.6% | 0.1% | 0.4% | 0.2% |
| ARIA-H (symptomatic) | 0.7% | 0.2% | 0.8% | 0.2% |

While most ARIA in CLARITY-AD and TRAILBLAZER-ALZ 2 was recorded as asymptomatic by site invesigators, there were 3% of participants on lecanemab and 6% of participants on donanemab who had symptoms. ARIA symptoms can include headache, confusion, seizure, or hemorrhagic stroke. In TRAILBLAZER-ALZ 2, 3.3% permanently discontinued donanemab treatment due to ARIA, and three cases of ARIA were fatal. In CLARITY-AD, fatal events and discontinuations were not reported separately for ARIA, but 6.9% permanently discontinued lecanemab treatment for any reason, including reasons unrelated to ARIA. The mortality rate was not statistically different in treatment compared with placebo in either trial (0.7% vs. 0.8% for lecanemab, and 1.9% vs. 1.0% for donanemab).

The presence of cerebral amyloid angiopathy (CAA)(29) or one or two copies of the *APOE* ε4 allele increases the risk of ARIA(29). In CLARITY-AD and TRAILBLAZER-ALZ2, the risk of ARIA was modified by *APOE* ε4 status (Table 3). There was a strong, graded relationship between ARIA risk and *APOE* status for both lecanemab and donanemab, with 40-50% of *APOE* homozygotes experiencing ARIA.

**Table 3.** Rates of ARIA and Mean CDR-SB Change According to *APOE* Genotype. ARIA risk and clinical efficacy according to *APOE* genotype in the CLARITY-AD (lecanemab) and TRAILBLAZER-ALZ 2 (donanemab) trials. Values are percent (for ARIA) or, for CDR-SB, the adjusted mean difference from placebo (Ref). In TRAILBLAZER-ALZ 2, only the subset of symptomatic ARIA that was considered “serious” (i.e., resulted in death, was life-threatening, required hospitalization, or caused persistent disability) was reported by *APOE* status. For ease of comparison, change in CDR-SB is shown for both trials; the interaction between *APOE* status and iADRS (the primary outcome in TRAILBLAZER-ALZ 2) was similar.

|  | **Lecanemab**<br>***APOE* ε4 status** |  |  | **Placebo**<br>***APOE* ε4 status** |  |  |
| --- | --- | --- | --- | --- | --- | --- |
|  | None<br>N=278 | Heterozygote<br>N=479 | Homozygote<br>N=141 | None<br>N=28<br>6 | Heterozygote<br>N=478 | Homozygote<br>N=133 |
| ARIA-E | 5.4% | 10.9% | 32.6% | 0.3% | 1.9% | 3.8% |
| Symptomatic<br>ARIA-E | 1.4% | 1.7% | 9.2% | 0% | 0% | 0% |
| ARIA-H | 11.9% | 14.0% | 39.0% | 4.2% | 8.6% | 21.1% |
| CDR-SB | -0.76<br>(-1.16, -0.35) | -0.51<br>(-0.79, -0.23) | +0.28<br>(-0.35, +0.88) | Ref | Ref | Ref |

|  | **Donanemab**<br>***APOE* ε4 status** |  |  | **Placebo**<br>***APOE* ε4 status** |  |  |
| --- | --- | --- | --- | --- | --- | --- |
|  | None<br>N=255 | Heterozygote<br>N=595 | Homozygote<br>N=143 | None<br>N=25<br>0 | Heterozygote<br>N=474 | Homozygote<br>N=146 |
| ARIA-E | 15.7% | 22.8% | 40.6% | 0.8% | 1.9% | 3.4% |
| Serious<br>ARIA-E | 0.4% | 1.8% | 2.8% | 0% | 0% | 0% |
| ARIA-H | 18.8% | 32.4% | 50.3% | 11.2% | 12.0% | 20.5% |
| Serious<br>ARIA-H | 0.4% | 0.2% | 1.4% | 0% | 0% | 0% |
| CDR-SB | -0.76<br>(-1.21, -0.31) | -0.73<br>(-1.06, -0.40) | -0.41<br>(-1.00, 0.19) | Ref | Ref | Ref |

Guidance on ARIA management is provided by the FDA package labels for lecanemab and donanemab, and from expert consensus recommendations for lecanemab(30). Management is based on the intersection of clinical symptom severity with radiological severity, according to provided definitions, and generally consists of extra MRI monitoring for asymptomatic radiologically mild ARIA, suspending doses until radiological resolution or stabilization for mild to moderate clinical symptoms and radiological signs, and permanently discontinuing drug for severe clinical symptoms or severe radiological signs. With this management, ARIA-E usually resolves within 4-8 weeks(29).

### Efficacy in subgroups

In the CLARITY-AD and TRAILBLAZER-ALZ 2 trials, effects were reported in some subgroups, including age, sex, AD stage (MCI vs dementia), race/ethnicity, and, for TRAILBLAZER-ALZ 2, tau level(3,4). Caution should be exercised when interpreting the results of these multiple post-hoc exploratory tests(31), which should be considered hypothesis-generating and not definitive. Adjusted mean differences in treatment vs placebo are reported here for the primary trial outcomes: for CLARITY-AD, the CDR-SB, in which *negative* differences favour treatment with lecanemab; and for TRAILBLAZER-AL2, the iADRS, in which case *positive* differences favour treatment with donanemab.

Across all primary and secondary end points, lecanemab and donanemab slowed decline on cognitive and functional composite scales significantly more than placebo for participants in the 65-74 years and ≥ 75 years age groups (supplemental figures S1(3) and E9(4), respectively). In the <65 year group, lecanemab and donanemab slowed decline but not significantly more than placebo although the confidence limits were wide (lecanemab -0.08 [95% CI -0.51 to +0.33]; donanemab low/medium tau population +2.09 [95% CI -3.36 to 7.66]; donanemab combined tau population +2.24 [95% CI -2.17 to +6.70]).

Efficacy was assessed according to participants’ sex in both trials. In CLARITY-AD, lecanemab was less efficacious, and not statistically significant, in females (-0.20 [95% CI -0.52 to +0.09], 12% slowing) compared with males (-0.73 [95% CI -1.01 to -0.42], 43% slowing). A similar lack of statistical significance in females was seen for multiple secondary outcomes including neuropsychological testing and activities of living. In contrast, in TRAILBLAZER-ALZ 2, donanemab had similar or greater effectiveness in females than males: +3.38 [95% CI +1.12 to +5.28]) vs +3.15 [95% CI 1.63 to 5.28] in the low/medium tau population, and +3.51 [95% CI +1.66 to 5.38]) vs +2.09 [95% CI -0.13 to +4.31] in the combined tau population.

The CLARITY-AD(3) and TRAILBLAZER-ALZ 2(4) trials included participants with MCI or dementia. In CLARITY-AD, lecanemab was efficacious for participants with MCI (-0.35 [95% CI -0.60 to -0.13, 41% slowing) and mild dementia (-0.62 [95% CI -1.06 to -0.18], 22% slowing)(3). In TRAILBLAZER-ALZ2, AD stage was classified based on Folstein MiniMental Status Exam as MCI (≥27), mild dementia (MMSE 20-26), and moderate dementia (MMSE <20)(4). In the low/medium tau population, donanemab was borderline efficacious in MCI (+2.92 [95% CI +0.04 to +1.63], 55% slowing) and efficacious in mild dementia (+ 2.54 [95% CI +0.93 to +4.25, 30% slowing) and moderate dementia (+5.51 [95% CI +2.24 to +8.87], 35% slowing). In the combined tau population, donanemab trended toward efficacy in MCI (+2.14 [95% CI -1.20 to +5.48], 39% slowing) and was efficacious in mild dementia (+2.25 [95% CI +0.54 to +4.00], 19% slowing) and moderate dementia (+3.70 [95% CI +0.84 to +6.70], 18% slowing). In TRAILBLAZER-AL 2, the efficacy of donanemab was weaker, and not statistically significant, in participants with high tau (+1.26 [95% CI -1.71 to +4.31]) than low/medium tau (+3.25 [95% CI +1.86 to +4.71]).These data raise the possibility that the effect of treatment, expressed as the percent slowing of progression, decreases as the AD stage worsens, with accumulating tau burden, from MCI to moderate dementia (although point estimates for the MCI stage are accompanied by wide confidence limits).

Both trials assessed efficacy according to the apolipoprotein E epsilon 4 (*APOE*4) allele status of participants (Table 3). Both lecanemab and donanemab were efficacious in the subgroups with no ε4 allele and *APOE* ε4 heterozygotes. However, in *APOE* ε4 homozygotes the effects were closer to the null and not statistically significant, but with wide confidence intervals (lecanemab: +0.28 [95% CI -0.35 to +0.88]; donanemab low/medium tau +1.91 [95% CI -1.40 to +5.32]; donanemab combined tau +1.01 [95% CI -2.37 to +4.36]). A dose-response relationship was seen in both trials, where effect sizes across all end points decreased with the number of ε4 allele copies.

While subgroup analyses were performed to assess efficacy of lecanemab and donanemab according to race and ethnicity, these analyses were considerably underpowered as non-Whites represented 23.2% and 8.5% of participants in the CLARITY-AD and TRAILBLAZER-ALZ 2 trials, respectively(3,4). For lecanemab, there was no evidence of different effects by race--classified as White, Asian, or Black—or ethnicity, classed as Hispanic or not Hispanic. In the TRAILBLAZER-ALZ 2 trial of donanemab, where race and ethnicity were categorized as in CLARITY-AD, the point estimate for Blacks favoured placebo rather than donanemab, but the confidence intervals were very wide, in the low/medium tau population (-2.68 [95% CI -10.69 to +5.41]) and the combined population (-2.35 [95% CI -11.64 to +6.80]). Additionally, the efficacy for Hispanics was not statistically significant confidence intervals in the low/medium tau population (-1.24 [95% CI -8.35 to +6.02]) and the combined population (+1.28 [95% CI -6.29 to +8.94]). Data were not available for North American Indigenous persons or subgroups of Asian ethnicities.

Subgroup analyses of ARIA risk were confined to the relationship with *APOE* genotype, which we reported in the section on ARIA. The trials did not provide risks of ARIA-E and ARIA-H according to age group, sex, or clinical stage of AD(3,4).

In summary, there were significant knowledge gaps with respect to potential efficacy in important subgroups. The potential decreased efficacy of lecanemab in female participants in the CLARITY-AD trial is concerning, considering AD affects more females than males around the globe. Future studies should investigate specifically the effects of sex and gender on efficacy and safety as these factors may have clinical importance in personalizing therapies for a patient. Additionally, future trials should recruit more diverse populations to increase confidence in the efficacy and safety of treatment across different ethnic groups.

Future studies should investigate efficacy and safety in patients with early-onset (< 65 years) and preclinical phases of AD (*i*.*e*., asymptomatic amyloid positive individuals). As well, the effects of anti-Aβ mAbs should also be assessed in the “old-old” (i.e., ≥ 85 years) and in the frail adults, considering the increasing incidence and prevalence of AD with aging and the increasing prevalence of comorbidity and polypharmacy with aging.

## CONSIDERATIONS FOR PROVIDING TREATMENT IN CLINICAL PRACTICE

### Patient selection in clinical practice

The selection criteria of CLARITY-AD and TRAILBLAZER-ALZ 2 were complex, with information collected at screening that is not part of usual clinical care (Table 4). Selection of patients for treatment in practice will require simpler, more pragmatic criteria. The FDA package labels for lecanemab and donanemab recommend that they are indicated for mild cognitive impairment or mild stage dementia due to AD (Table 4). A group of experts has provided consensus recommendations for selection of patients for treatment in routine practice(30).

**Table 4.**
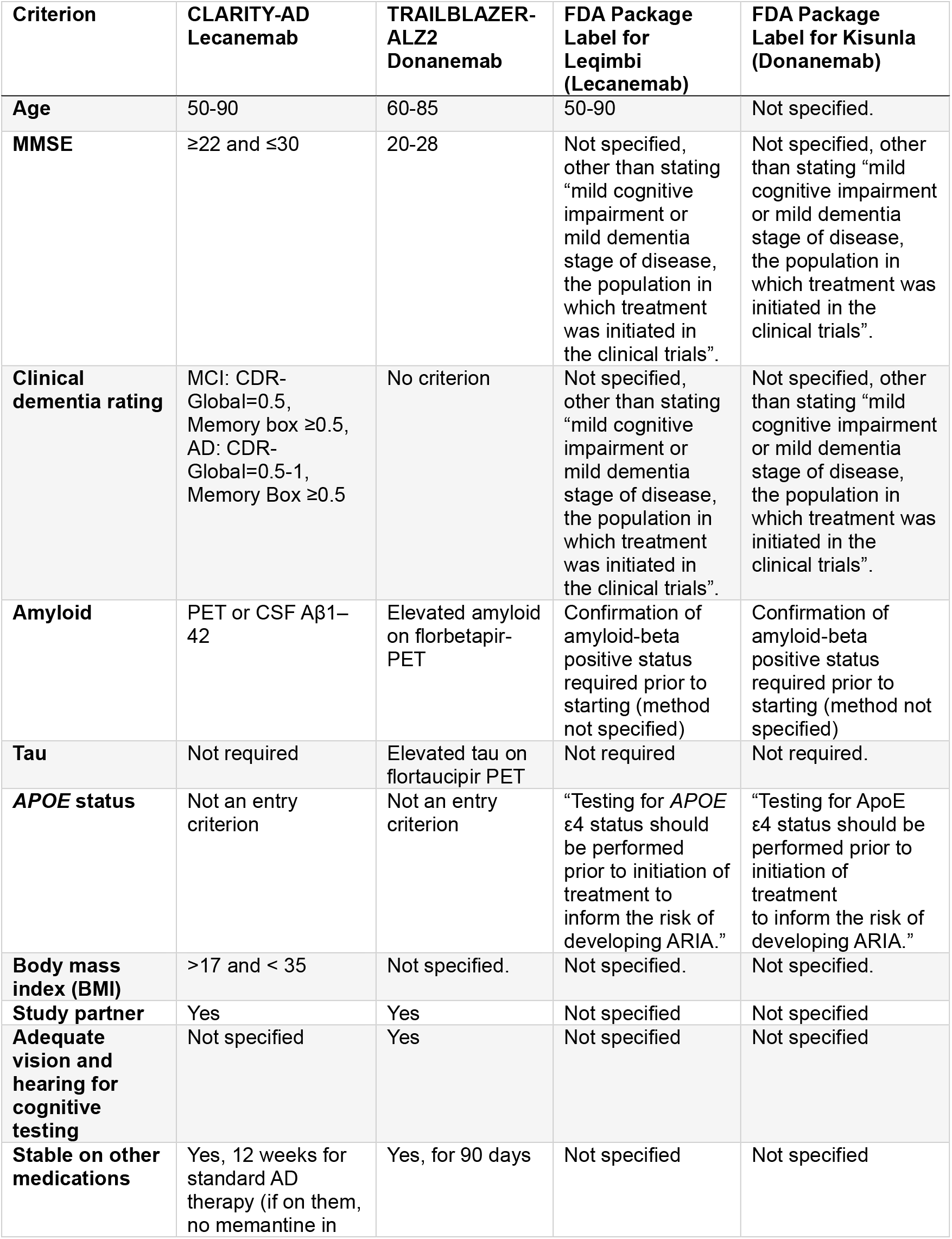
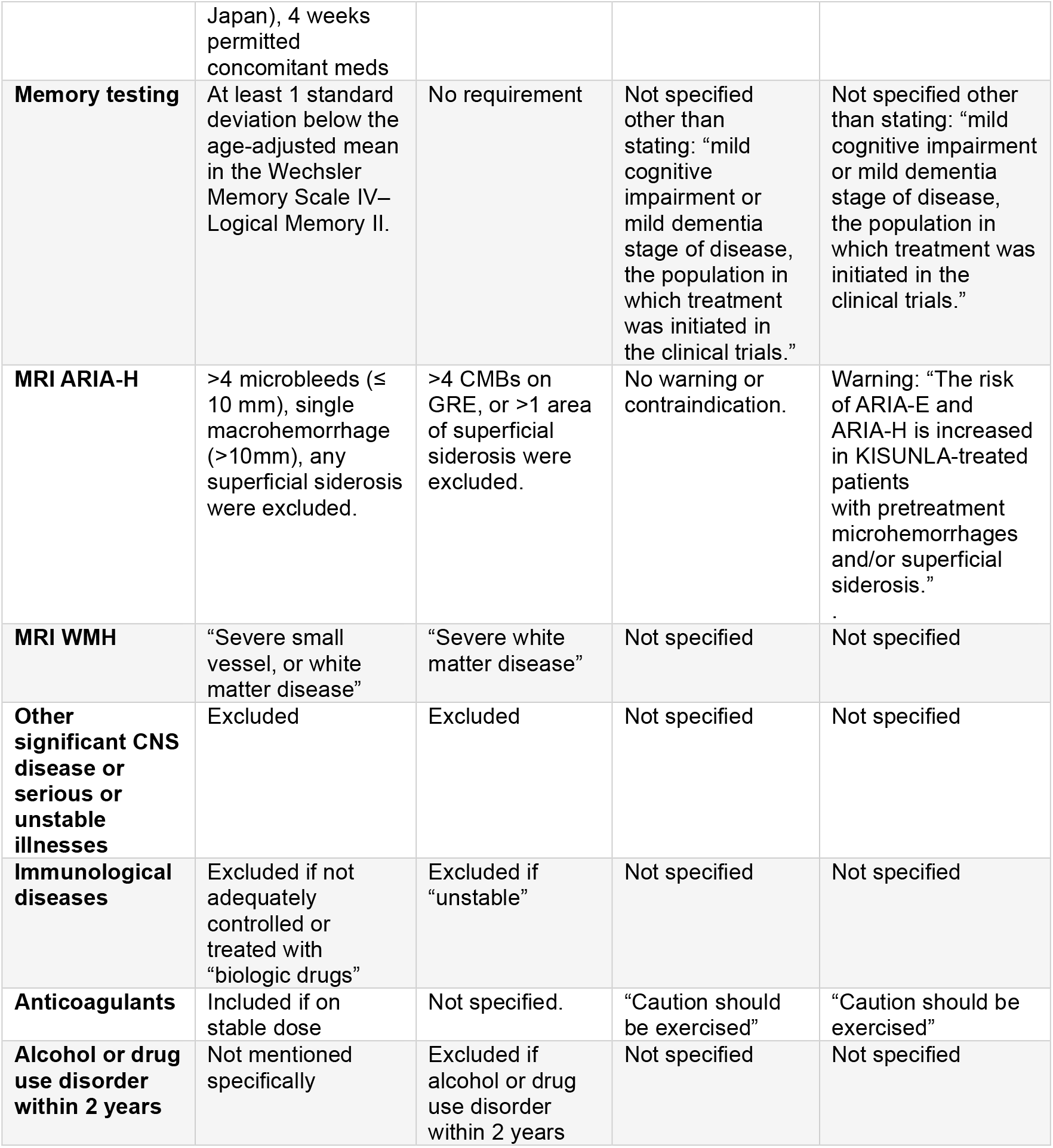
Patient selection criteria used in the clinical trials and on the FDA package labels.

| **Criterion** | **CLARITY-AD**<br>**Lecanemab** | **TRAILBLAZER-**<br>**ALZ2**<br>**Donanemab** | **FDA Package**<br>**Label for**<br>**Leqimbi**<br>**(Lecanemab)** | **FDA Package**<br>**Label for Kisunla**<br>**(Donanemab)** |
| --- | --- | --- | --- | --- |
| **Age** | 50-90 | 60-85 | 50-90 | Not specified. |
| **MMSE** | ≥22 and ≤30 | 20-28 | Not specified, other than stating “mild cognitive impairment or mild dementia stage of disease, the population in which treatment was initiated in the clinical trials”. | Not specified, other than stating “mild cognitive impairment or mild dementia stage of disease, the population in which treatment was initiated in the clinical trials”. |
| **Clinical dementia rating** | MCI: CDR-Global=0.5, Memory box ≥0.5, AD: CDR-Global=0.5-1, Memory Box ≥0.5 | No criterion | Not specified, other than stating “mild cognitive impairment or mild dementia stage of disease, the population in which treatment was initiated in the clinical trials”. | Not specified, other than stating “mild cognitive impairment or mild dementia stage of disease, the population in which treatment was initiated in the clinical trials”. |
| **Amyloid** | PET or CSF Aβ1–42 | Elevated amyloid on florbetapir-PET | Confirmation of amyloid-beta positive status required prior to starting (method not specified) | Confirmation of amyloid-beta positive status required prior to starting (method not specified) |
| **Tau** | Not required | Elevated tau on flortaucipir PET | Not required | Not required. |
| **APOE status** | Not an entry criterion | Not an entry criterion | “Testing for APOE ε4 status should be performed prior to initiation of treatment to inform the risk of developing ARIA.” | “Testing for ApoE ε4 status should be performed prior to initiation of treatment to inform the risk of developing ARIA.” |
| **Body mass index (BMI)** | >17 and < 35 | Not specified. | Not specified. | Not specified. |
| **Study partner** | Yes | Yes | Not specified | Not specified |
| **Adequate vision and hearing for cognitive testing** | Not specified | Yes | Not specified | Not specified |
| **Stable on other medications** | Yes, 12 weeks for standard AD therapy (if on them, no memantine in | Yes, for 90 days | Not specified | Not specified |
|  | Japan), 4 weeks permitted concomitant meds |  |  |  |
| **Memory testing** | At least 1 standard deviation below the age-adjusted mean in the Wechsler Memory Scale IV–Logical Memory II. | No requirement | Not specified other than stating: “mild cognitive impairment or mild dementia stage of disease, the population in which treatment was initiated in the clinical trials.” | Not specified other than stating: “mild cognitive impairment or mild dementia stage of disease, the population in which treatment was initiated in the clinical trials.” |
| **MRI ARIA-H** | >4 microbleeds ($\leq 10$ mm), single macrohemorrhage (>10mm), any superficial siderosis were excluded. | >4 CMBs on GRE, or >1 area of superficial siderosis were excluded. | No warning or contraindication. | Warning: “The risk of ARIA-E and ARIA-H is increased in KISUNLA-treated patients with pretreatment microhemorrhages and/or superficial siderosis.” |
| **MRI WMH** | “Severe small vessel, or white matter disease” | “Severe white matter disease” | Not specified | Not specified |
| **Other significant CNS disease or serious or unstable illnesses** | Excluded | Excluded | Not specified | Not specified |
| **Immunological diseases** | Excluded if not adequately controlled or treated with “biologic drugs” | Excluded if “unstable” | Not specified | Not specified |
| **Anticoagulants** | Included if on stable dose | Not specified. | “Caution should be exercised” | “Caution should be exercised” |
| **Alcohol or drug use disorder within 2 years** | Not mentioned specifically | Excluded if alcohol or drug use disorder within 2 years | Not specified | Not specified |

There is consensus that *APOE* genotype testing, which is not currently part of clinical practice in Canada, should be done prior to treatment to inform the risk of ARIA. The risk of ARIA increases with each additional copy of the *APOE* ε4 allele. *APOE* ε4 homozygotes are at especially high risk for ARIA, and probably derive less benefit from treatment (Table 4; also see the sections on ARIA and Efficacy in Subgroups). The FDA package labels for lecanemab and donanemab recommend that *APOE* ε4 status should be ascertained prior to treatment, and that ARIA risk should be discussed with patients. Notably, the UK MHRP for lecanemab excludes individuals who are *APOE* ε4 homozygotes. The CCNA recommends that, in obtaining consent for treatment with anti-Aβ mAbs, clinicians should cite the higher risk and potential for less benefit in persons who are *APOE* ε4 homozygotes.

Patients taking anticoagulants or given thrombolytics may be at risk for intracranial hemorrhage, based on limited data. Media reports have indicated that two trial participants taking lecanemab had fatal intracranial hemorrhages while taking anticoagulants(32), and one participant taking lecanemab had intracranial hemorrhage after being treated with thrombolysis for acute ischemic stroke(33). Data presented at a scientific conference, but not yet published in a peer-reviewed journal, indicated that 2/140 participants on anticoagulants died with concurrent macrohemorrhage while taking lecanemab compared with 0/74 patients on anticoagulants taking placebo(30). The FDA package labels for lecenamab and donanemab recommend “caution” when treating patients on anticoagulants. Many of the ongoing trials of the anti-Aβ mAbs are excluding patients on anticoagulants. So far, no safety concerns have arisen in patients taking antiplatelet drugs such as aspirin; however, there are insufficient data on patients taking multiple antiplatelet drugs. In accordance with an expert consensus group(30), the CCNA recommends against treatment of patients on anticoagulants.

Patients with variant AD phenotypes (including posterior cortical atrophy, logopenic aphasia, or frontal behavioural variant) could have early-stage AD but not satisfy CLARITY-AD or TRAILBLAZER-ALZ 2 cognitive testing criteria due to the nature of their symptoms, including confounding of testing by visual impairment or aphasia. Whether these patients can be treated safely and effectively is not clear. If treatment of these patients is contemplated, it will be essential to prove that amyloid-beta, the target for treatment, is present in the brain, and that functional impairment is mild, indicating mild AD analogous to the CLARITY-AD and TRAILBLAZER-ALZ 2 inclusion criteria. In patients with posterior cortical atrophy or logopenic aphasia, the MMSE may be confounded by disproportionate visuospatial or language dysfunction, and cut-offs used in the trials may not be appropriate.

Patients with severe WMH or multiple infarcts, suggesting a vascular contribution to cognitive decline, were excluded from the trials. The CCNA Work Group suggested that these patients should be excluded from treatment in routine clinical practice, as well. Whether treatment benefits patients with significant cerebrovascular disease as well as a positive amyloid-beta biomarker is an important question that warrants further study in future clinical trials.

In the TRAILBLAZER-ALZ 2 trial of donanemab, elevated tau-PET was required for eligibility and recruitment was stratified by tau level(4). However, the FDA label for donanemab does not require testing for tau, and the CCNA Work Group similarly recommends that assessment of tau is not necessary prior to treatment.

### Diagnostic confirmation of AD

Enrollment in CLARITY-AD or TRIALBLAZER-ALZ 2 required biomarker confirmation of amyloid-beta in the brain(3,4). The CCNA Work Group agreed that this should be retained as a criterion for receiving anti-Aβ mAbs, given prior evidence that the rate of false positive AD diagnosis is unacceptably high (approximately 25%(34)) without AD biomarker testing. The best validated diagnostic tests for amyloid-beta are amyloid-PET (in the trials, florbetaben(3), flutemetamol(3), or florbetapir(3,4) ligands were used) and CSF Aβ42 measured and reported as a ratio with either tau proteoforms (total-tau [t-tau] or phosphorylated-tau [p-tau]) or Aβ40. However, AD biomarker testing is not done routinely in patients suspected of AD in Canada. The Canadian Consensus Conference on Diagnosis and Treatment of Dementia (CCCDTD) recommends that amyloid-PET or CSF AD biomarker testing are not needed for routine diagnosis, but may be useful for patients in whom the underlying pathological process is not clear despite specialist evaluation(35). The use of amyloid imaging should follow guidelines from the Specialized Task Force on Amyloid Imaging in Canada(36).

The CCNA Work Group recommends that amyloid-PET is the preferred method for providing diagnostic confirmation of brain amyloid-beta. The anti-Aβ mAbs lecanemab and donanemab have been validated to reduce amyloid-PET signal(3,4). In general, across all anti-Aβ mAbs the degree of amyloid PET signal reduction correlates with the degree of clinical benefit(37). Furthermore, in the TRAILBLAZER-ALZ2 trial, treatment with donanemab was stopped if the amyloid-PET signal was normalized(4). The CCNA Work Group recommends that the SNMMI Procedure Standard/EANM Practice Guideline for Amyloid PET Imaging of the Brain(38) should be used as a guide to acquiring, processing, and interpreting those studies. The use of amyloid imaging should follow guidelines from the Specialized Task Force on Amyloid Imaging in Canada(36).

A challenge to using PET for AD diagnosis in Canada is the limited and variable access across provinces. There are only 45 PET cameras in Canada, with 24 in Quebec and 12 in Ontario(39). Florbetaben is the sole imaging agent for amyloid-beta that is used clinically in Canada. but its production is confined to Quebec and Ontario. Although cyclotrons are present in other regions, enabling potential synthesis at these sites, scanning capacity is restricted.

The current alternative to amyloid PET is lumbar puncture (LP) for CSF analysis. In AD, the concentration of abeta peptide 1-42 (Aβ42) in CSF is reduced(40). The highest diagnostic performance for CSF biomarkers is achieved when Aβ42 is reported as a ratio to Aβ40, p-tau or t-tau. AD CSF biomarkers are highly concordant with amyloid-PET and the gold standard, neuropathological evaluation(41,42). Although well validated for AD diagnosis, there are fewer data on the responsiveness of CSF AD biomarkers to anti-Aβ mAbs; therefore, it is not yet clear whether repeat CSF analyses can be used to monitor therapy. In TRAILBLAZER-AL2, thresholds to cease donanemab therapy were based on amyloid-PET, not CSF measures.

A prior model estimated that only 1.15% of Canadian patients with mild dementia or MCI due to AD had access to AD biomarker testing, assuming that half would have amyloid PET and half would have CSF testing(7). Switching to an all-CSF diagnostic strategy would increase the capacity by 46,000 per year; however, it was not clear whether the model inputs included variables related to LP availability(7). Capacity for performing LPs is limited and is centered in urban specialty practices. Typically, LP is within the scope of practice for neurologists and anaesthesiologists, but not geriatricians or geriatric psychiatrists. A survey of the CCNA membership indicated that CSF testing is available and reimbursed by provincial health authorities in British Columbia for patients meeting appropriate use guidelines(41), and in specialty dementia clinics in Alberta, Ontario, and Quebec. Currently, a clinical laboratory in British Columbia performs CSF testing with Health Canada-approved testing kits; elsewhere, testing must be sent out of province, and in some provinces special approvals may be required.

The accuracy of plasma markers of AD is approaching that of CSF or amyloid-PET(43), offering the potential for easier access to diagnostic testing. The highest diagnostic accuracy is not for plasma Aβ, but rather for tau proteoforms phosphorylated at specific sites. Current data suggest that plasma ptau-217 has the best sensitivity and specificity for AD, including in early stages, and can discriminate it from other causes of neurodegeneration including non-AD tauopathies(44,45). However, more data are needed on the impact of assay types, clinical settings, polytherapy, and multiple comorbidities on test performance(43,46). Currently, there are no Health Canada licensed blood-based diagnostic kits for AD biomarker testing. Multiple manufacturers have created plasma-based test kits, and related trials (outside of Canada) are in progress with the aim of obtaining regulatory approval. Current unknowns are how and when provinces will build capacity for plasma testing, whether a centralized versus distributed model will be used. Additionally, appropriate use guidelines and reporting standards will need to be developed and implemented(47).

### Neuroimaging protocols

To use the anti-Aβ mAbs in clinical practice will require pre-treatment and follow-up MRI and, ideally, amyloid-PET, although in many regions amyloid-beta may have to be diagnosed by CSF analysis, instead. In the Canadian context, requirements for more MRI and PET would place a significant burden on radiological and nuclear medicine resources. The availability of MR and PET imaging in Canada varies greatly between provinces. Additional research and planning are needed to clarify MRI and PET capacity, with respect to the number of treatment candidates in any given region.

In the CLARITY-AD and TRAILBLAZER-ALZ 2 trials, follow-up MRIs were required to screen for the presence of ARIA-E and ARIA-H; however, the exact frequency and timing varied by trial (Figure 1). Unfortunately, MRI technical parameters were not specified in sufficient detail in the trial publications and supplemental trial protocol documents to reproduce the drug-specific MRI protocol in routine practice, with incomplete details on the MRI field strength, slice thickness, and sequence types. It appears likely that the trial protocols followed 2011 consensus recommendations for ARIA screening, including minimum field strength 1.5T, maximum slice thickness of 5 mm without any specification on slice gaps, and use of the gradient recalled echo (GRE) sequence as it was “presently available on any scanner worldwide”)(28).

A challenge with using the trial protocols in routine practice is that imaging protocols have advanced, with (28),increasing adoption of more sensitive techniques including use of higher field strength (3T) and newer methods such as susceptibility imaging that can detect hemorrhage more readily. Susceptibility imaging at 3T can detect up to twice as many microbleeds, on average, as on GRE at 1.5 Tesla field strength(48). This creates a dilemma, as t is uncertain whether the trial thresholds for hemorrhagic lesions (>4 microbleeds or any [CLARITY-AD(3)] or more than one [TRAILBLAZER-ALZ 2(4)] area of superficial siderosis) are applicable when using higher sensitivity imaging.. More research is needed on the comparability of modern compared with older generation MRI imaging to predict and diagnose ARIA-H and ARIA-E. However, the Work Group recommends that, because of their superior overall diagnostic accuracy, dementia imaging protocols should continue to use the best sequences available at that site, including susceptibility imaging (Table S1).

MRI was used in the trials to monitor the risk for ARIA (Figure 1) (3,4). Because a diagnostic image has already been obtained, a monitoring MRI protocol could potentially be much shorter: a fluid attenuated inversion recovery (FLAIR) for ARIA-E and a hemorrhage sensitive sequence, preferably susceptibility imaging, for ARIA-H (Supplemental Table S1). This acquisition time for this short duration monitoring protocol could as little as 10 minutes. To maximize comparability across scan sessions, the same protocol, and ideally the same scanner, should be used for each individual patient.

Adoption of the anti-Aβ mAbs would also require changes in radiological reporting. Determining eligibility for therapy and grading the severity of ARIA events requires reporting of the axial diameter of areas of ARIA-E and the exact count of the number of hemorrhagic lesions. Radiologists would need to become familiar with accepted grading schemes, terms, and definitions of radiological manifestations of ARIA. The use of standard reporting templates, which could be embedded within electronic health records, and continuing medical education modules could help achieve greater standardization and quality.

### Organizing clinical care including infusions

The anti-Aβ mAbs present specific challenges to the organization of clinical care. Lecanemab is administered intravenously over one hour every two weeks, while donanemab is administered intravenously over one hour every four weeks(3,4). Therefore, administering these treatments requires infusion service capacity, either in in a hospital setting, out-patient infusion facility or, potentially, via home nursing visits. A list of equipment, supplies, and human resources required to administer lecanemab and donanemab is shown in Table S2.

Lecanemab and donanemab can elicit infusion-related reactions, occurring at the time of infusion, or up to several hours after the infusion, causing fever, chills, headache, rash, nausea, vomiting, abdominal discomfort, and elevated blood pressure (3,4). The rate of any infusion reactions was higher for lecanemab than donanemab (26.5% versus 8.7%) but so was the rate of placebo infusion reactions (7.4% versus 0.5%), suggesting that some of the difference may have been due to more sensitive ascertainment of adverse events. In contrast, the rate of serious infusion reactions was similar (1.2% for lecanemab versus 0.4% for donanemab). If a reaction occurs, the infusion should be stopped, and the patient may be treated with diphenhydramine and acetaminophen, or with oral dexamethasone or oral methylprednisolone when marked symptoms are present(30).

Because expertise in AD diagnostic testing and ongoing monitoring for ARIA are required, lecanemab and donanemab should be prescribed by a dementia specialist (*e*.*g*., neurologist, geriatrician, geriatric psychiatrist) or a specialized family physician with extensive experience in the diagnosis and treatment of cognitive disorders, such as AD. A list of equipment, supplies, and human resources required to administer lecanemab and donanemab is shown in Supplemental Table S2. The care team should include a nurse or physician assistant who can assist the prescribing physician with logistical support surrounding ordering and receiving the results of baseline amyloid biomarkers and MRI, as well as ordering and receiving results of monitoring MRIs (i.e. assessing for emerging ARIA), in advance of continuing intravenous therapy. Involvement of a specialist would likely be required for the full duration of lecanemab and donanemab therapy; however, models of care could be developed whereby centres of expertise provide remote support via telemedicine. Because infusion-related reactions are most likely to occur within the first 9-13 weeks(30), hybrid models can be considered which may involve infusions given at centres of expertise for the initial period of treatment, followed by maintenance infusions given closer to a patient’s primary residence. Models of care would need to be adapted to the organization of health regions, with their specific geographical and social features.

A subcutaneous formulation of lecanemab is being tested in clinical trials, which would allow at-home injection of lecanemab on a once-weekly basis. The availability of a self-injection device partially mitigates the need for home or clinic-based nursing care, but some support will still be necessary for education and support, including patients and caregivers who are unable to self-inject on their own.

One of the most important unanswered questions in therapy is when infusions should cease. In the TRAILBLAZER-ALZ II trial of donanemab, treatment was stopped if amyloid-PET signal normalized (which was the case in 29.7% of participants at 6 months and 76.4% at 12 months)(4) while in CLARITY-AD(3), and in TRAILBLAZER-ALZ II participants without amyloid clearance, infusions continued until 18 months. Currently, there are no published data on the efficacy of lecanemab or donanemab beyond 18 months, although evidence will eventually be available from open-label extensions of both trials.

It is currently unknown whether treatment should be continued into the more advanced stages of dementia, when tau and vascular amyloid deposition will have accumulated to a greater degree. Many experts believe that higher tau may be associated with less clinical response to amyloid removal(4), and the presence of vascular amyloid (*i*.*e*., cerebral amyloid angiopathy) increases the risk of ARIA(29). Thus, it is possible that the balance of benefits and risks is less favourable in later stage AD. However, in the absence of clear consensus guidelines it may be difficult for clinicians to stop therapy in patients desiring to continue treatment.

If amyloid is cleared, it is uncertain whether amyloid accumulates and how rapidly. If amyloid re-accumulates, it is not clear whether treatment should be re-initiated and what biomarker or clinical threshold should prompt re-initiation. These questions are being examined in ongoing studies.

Creation of a pan-Canadian longitudinal AD treatment registry will be essential to obtaining short- and long-term safety and efficacy data in the Canadian context. Such data may enable selection and expedited assessment of subjects who are most likely to benefit from treatment.

## IMPLICATIONS FOR DEMENTIA SYSTEMS OF CARE

### Role of primary care and equitable access to treatment

In Canada, primary care will have a critical role in identifying potential candidates for anti-Aβ mAbs and helping patients to understand potential benefits and harms of treatment.

Currently, primary care clinicians face many well-documented challenges in the assessment and timely accurate diagnosis of early-stage AD(49,50). These include lack of time, knowledge and training, inadequate remuneration, and in most provinces, mandatory reporting of potentially unfit drivers to transportation authorities which can negatively affect clinician-patient relationships. Diagnosis of cognitively impaired but non-demented individuals (*i*.*e*., MCI) has not been a priority in primary care(51), possibly because there are no currently approved pharmacological therapies. Further compounding access to timely diagnoses is the often-lengthy wait times for specialist consultation across many parts of Canada.

If anti-Aβ mAbs become available, it will be important for primary care practitioners to identify potentially eligible patients with cognitive disorders and to refer these persons along local care pathways to access treatment. Validated rapid screening tools, such as the Mini-Cog and AD-8, may be used to screen for those in need of more a more detailed cognitive assessment to establish a diagnosis(52). Education outlining local care pathways and potential benefits and harms of anti-Aβ mAbs for eligible patients can help primary care practitioners to knowledgeably counsel patients and families and to initiate appropriate referrals for accurate diagnoses and potential treatment. However, there are insufficient specialists in dementia care and many rural and remote communities have limited access(7). Referrals of patients who have not been fully evaluated in primary care may overwhelm specialist center waiting lists and further delay access.

In all Canadian provinces, family physicians have the primary responsibility to diagnose and manage patients with dementia, reserving specialist referrals for a minority of more complex cases. Some provinces have implemented special recommendations and programs to support family physicians, such as the Quebec Alzheimer Plan(53). To implement the use of anti-Aβ mAbs in Canada, these programs will need to be enhanced and to accommodate recommendations for anti-Aβ mAbs screening and use.

The College of Family Physicians of Canada offers a Certificate of Added Competence in the Care of the Elderly. While not exclusively focused on dementia, this certification program could be a vehicle for creating a cadre of family physicians with special expertise in screening for eligibility for anti-Aβ mAbs, and perhaps for administering them.

Adoption of new innovative care models may be needed to address these significant challenges. For example, the Multi-specialty INterprofessional Team (MINT) Memory Clinics, which are now located in over 100 sites across 6 provinces, integrate collaborative partnerships between primary care and specialist care(54,55). Through standardized nationally-accredited training for family physicians and multidisciplinary teams in primary care, the MINT Memory Clinic model has demonstrated better health outcomes(56), better experience of care for patients and caregivers and for healthcare providers(57), service to rural and marginalized populations(55), and lower healthcare costs(58).

The introduction of a new, complex, and expensive therapy that requires special competence for delivery has the potential to exacerbate disparities in care for populations that are currently underserved, including non-White, Indigenous, Lesbian, Gay, Bisexual, Transgender, Queer or Questioning, Two-Spirit, lower socioeconomic status, immigrant, and rural populations(59). The reduced access of rural patients to specialty care(60,61) must be addressed. Innovative programs for remote access to memory clinics, such as those being developed by Saskatchwan’s Rural Dementia Action Research team, may be one way to increase access(62).

### Projected numbers of eligible patients

The number of eligible patients will greatly affect the capacity of Canadian dementia systems to deliver anti-Aβ mAbs. Analyses of system preparedness have suggested that there is little reserve capacity within the system, with challenges that include access to AD biomarker testing, access to specialist care, and MRI wait list times.(6,7,9,10)

A simulation model for Canada predicted that 382,000 Canadians would be on the waitlist for eligibility assessment for anti-Aβ mAbs, but did not estimate the number of Canadians that would meet criteria for treatment after assessment(7). A limitation of the model was that it assumed that patients with primary care would be screened for MCI or dementia by their family physician, which is currently not recommended in primary care practice(63) and seems unrealistic to implement.

A report sponsored by Biogen and conducted by the RAND Corporation estimated that the number of persons over age 50 with MCI in Canada in 2020 was 1.75 million, of whom half would be positive for AD biomarkers(6). Using a simulation model, they estimated that Canada-wide population screening would result in 200,000 Canadians with MCI who were eligible for treatment and would seek treatment. However, there were many limitations to this model. Epidemiological studies show that the prevalence of MCI in older populations can vary by 16-fold (from 2.5% to 41%)(64) depending on which thresholds are used for cognitive test scores and how cognitive symptoms are elicited. Additionally, the RAND simulation model assumed that 80% of Canadians would agree to be screened and 50% of those who screened positive would see a specialist(6). These estimates are probably generous, as a recent randomized trial of dementia screening found that most participants (66%) who screened positive subsequently declined to see a specialist, even though it was part of the study protocol(65). This suggests that most elderly prefer not to be screened; however, it is not known whether the potential to be treated with a disease-modifying treatment, albeit one with partial effect, would increase their enthusiasm. Given the unclear prevalence of MCI and dementia due to AD, the uncertain validity and acceptability of screening methods, and the lack of data on long-term health benefits and cost effectiveness of anti-Aβ mAbs, the CCNA does not recommend population screening for anti-Aβ mAbs eligibility.

Based on current practice patterns, the number of patients eligible for treatment is likely to be much lower than these pharmaceutical company projection because of under-recognition, the presence of contraindications and, potentially, patient choice.

There is under-recognition of dementia and MCI in current practice, that will limit referrals for treatment. A systematic review estimated that only 38% of people in the community with dementia had a diagnosis in their medical record(49), and a study of U.S. Medicare data found that only 8% of expected MCI cases were diagnosed(51). With a new treatment, rates of diagnosis may improve, but this would probably take time.

Based on the complex selection criteria (Table 4), many patients will not be eligible due to medical comorbidities and other contraindications. An analysis of the population-based Mayo Clinic Study on Aging found that only 8% of persons with MCI or mild dementia and positive amyloid-PET met the other criteria for the CLARITY-AD trial(66). MRI findings of cerebrovascular disease, cardiac conditions, and recent active cancer were common reasons for exclusion(66). Relaxing the trial criteria by including patients with MCI regardless of cognitive test score thresholds resulted in 17.4% meeting criteria for treatment(66). A population-based study from Sweden found that only 12/30 patients with biomarker-positive AD met criteria for treatment with lecanemab(67). A study of UK community memory clinics found that 71% of patients had a diagnosis of possible AD and that only 32% had no medical or imaging contraindications to anti-Aβ mAbs and thus were eligible for AD biomarker testing(68). A study using Alberta administrative data found that at least 50% of persons with dementia would be ineligible based on medical comorbidities alone, before considering neuroimaging findings or AD biomarker results(69). Another study from an Alberta memory clinic found that only 23-34% of patients referred for MCI or AD would need amyloid-beta testing to determine eligibility for anti-Aβ mAbs; the rest could be excluded because they did not meet trial eligibility criteria due to cognitive test result criteria, comorbidities, or neuroimaging criteria(70).

An important unknown is how many eligible patients would choose treatment if it were offered. Lecanemab and donanemab are intensive, time-consuming treatments that require frequent healthcare visits, multiple MRI scans (Figure 1), and potentially an LP. Given this burden, it seems likely that some patients with not choose to have treatment, particularly if they are frail or have other comorbidities. Currently, there is little research on patient preferences for treatment.

Research in the Canadian context is needed to provide estimates of eligible patients in Canada, which are critically important for resource allocation. Canadian national estimates of the prevalence and incidence of dementia are provided by the Public Health Agency of Canada (PHAC), based on administrative health data(71), and Alzheimer Society of Canada. However, these data sources are not suited to directly estimate the number of Canadians eligible for treatment with anti-Aβ mAbs because dementia stage is not captured, MCI is not estimated, and there are only limited data on comorbidities that would preclude treatment.

In Canada, analyses of inclusion and exclusion criteria for treatment could be undertaken in population-based studies such as the Canadian Community Health Survey(72) and the Canadian Longitudinal Study on Aging(73), and from multicenter clinic-based cohorts such as the Comprehensive Assessment of Neurodegeneration and Dementia Study(74). However, currently there is no population-based study in Canada that contains all the information, including amyloid-beta biomarker status, needed to determine eligibility for anti-Aβ mAbs. Building capacity for dementia surveillance in Canada, including amyloid-beta biomarkers and neuroimaging, should be a priority.

## CONCLUSIONS

Lecanemab and donanemab are the first drugs to robustly lower cerebral amyloid-beta with partial effects on the rate of cognitive and functional decline, the clinical value of which continues to be debated. The drugs have been approved by some countries, including the US FDA, but not others. They are currently under review by Health Canada, which approves drugs for marketing in Canada, and the Canadian Drug Agency, which will issue a report that includes a cost effectiveness analysis.

Our review found strong evidence from phase 3 trials that lecanemab and donanemab slow the relative rate of decline on combined cognitive/functional scales by 27.1% and 22.3%, respectively, over an 18 month period. Effects on secondary outcomes were consistent with the primary outcomes. ARIA side effects were found radiologically in 21.5% and 36.8%, and were symptomatic in 2.8% and 6.1%. *APOE* ε4 homozygotes had a much higher risk of ARIA (close to 50%) and may not have derived clinical benefit, although conclusions about the efficacy in *APOE* ε4 homozygotes are limited by the post-hoc exploratory nature of these subgroup analyses, with relatively small numbers of participants and wide confidence intervals.

Implementing the anti-Aβ mAbs in Canada would require substantial changes in the organization of dementia care. Five major barriers stand out. First, there is under-diagnosis of early stage AD in primary care(49). Second, there is a lack of access to specialist care, which contributes to the problem of under-diagnosis. The current number of specialists is insufficient to manage the patients that might be eligible for treatment(7). Third, AD biomarker testing is limited to a small number of specialists in limited geographic regions. Increasing its availability would require investing in more capacity for amyloid-PET and CSF testing and, once validated and Health Canada-approved kits are available, plasma testing. Fourth, the MRIs required for monitoring and managing ARIA will place a burden on radiological services, with many Canadian jurisdictions are already experiencing long wait list times(75). Fifth, there would need to be systems of care for providing intravenous infusions.

There are many unanswered questions about the use of these therapies and their impact on the Canadian health system, deserving of future research. A list of some of the most important ones, generated by our Work Groups, is shown in Table 5. For individuals receiving therapy, it is not clear when therapy should stop, and, if amyloid-beta has been cleared, how long it takes to reaccumulate. Work will be needed to define the treatment eligible population in Canada, and patient and caregiver preferences for treatment. Advances in plasma testing may allow greater access to AD biomarkers. The cost effectiveness of therapy in Canada is not known, but will be explored by the Canadian Drug Agency. We did not summarize international data on cost effectiveness, because Canadian costs will differ. However, some studies are beginning to emerge(76-79). A company-sponsored study found that lecanemab was cost effective at a willingness-to-pay threshold of $100,000 per quality-adjusted life-year gained(76), but an academic analysis suggested that lecanemab was over-priced for the value of the clinical benefits(79).

**Table 5.**
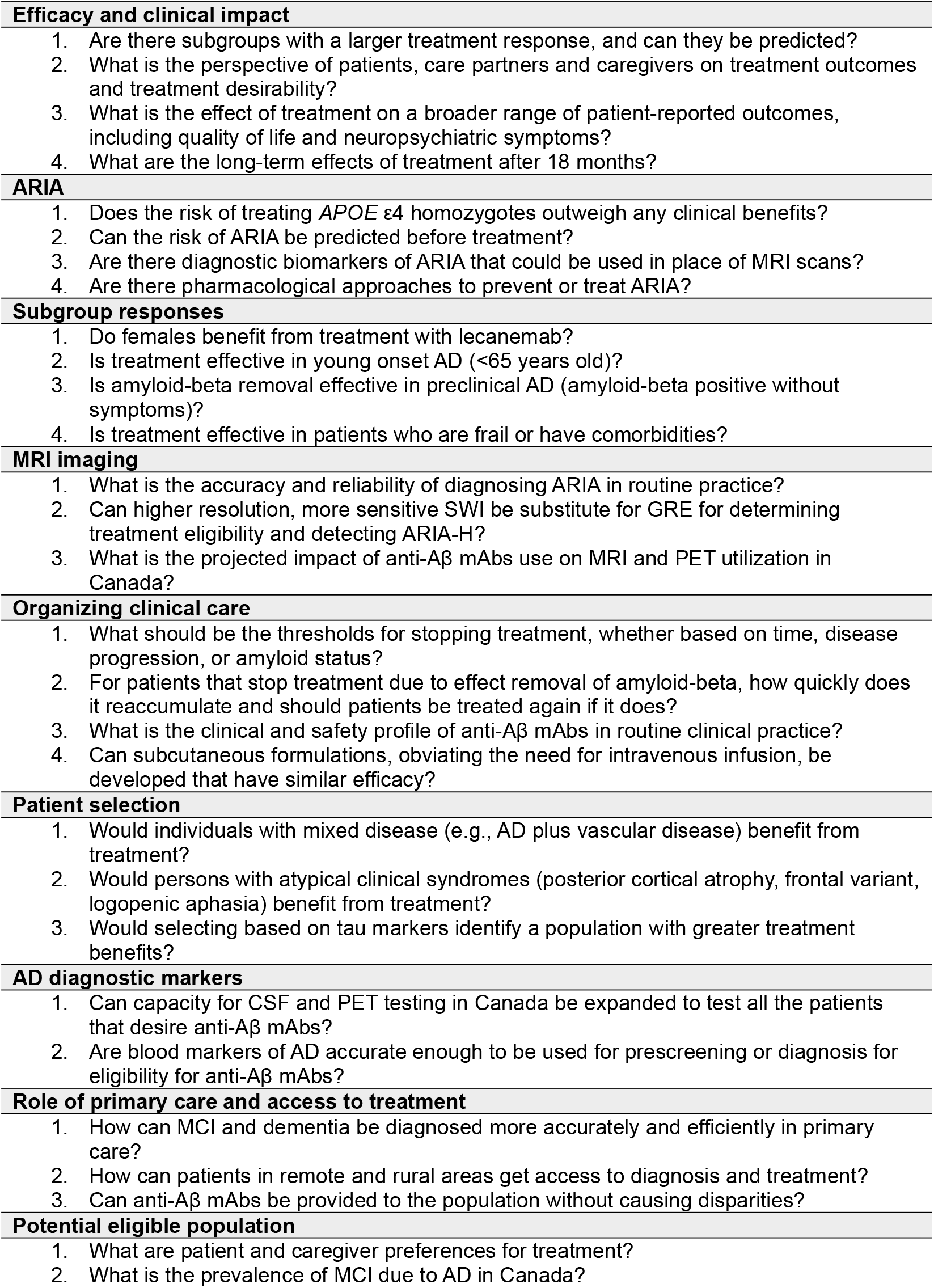
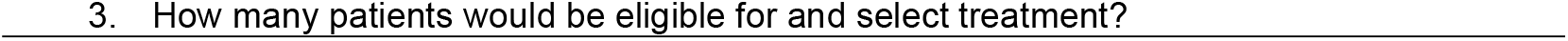
Research questions.

In conclusion, the discovery that AD outcomes can be improved by effectively removing amyloid-beta is a landmark development, opening the door to new treatment strategies for this common, dreaded disease. Future drugs in this class may, hopefully, address some of the limitations of lecanemab and donanemab, including the need for intravenous infusion and the risk of ARIA. The findings of CLARITY-AD and TRAILBLAZER-ALZ 2 suggest that amyloid-beta removal will probably be a component of AD disease-modifying treatment, but to fully halt progression more effective drugs or combination approaches will still need to be discovered.

## Data Availability

This study uses meta-data from previously published papers. All data produced in the present work are contained in the manuscript.

## ACKNOWLEDGEMENTS

EES (Chair), NP, HF, MB, and HC comprised the Steering Committee. AG, AH-B, PD, and AF Chaired Work Groups. RC and MM provided internal independent peer review. The other authors participated as members of Work Groups.

## List of Tables and Figures

Table S1. MRI protocol recommendations

Table S2. Equipment, supplies, and human resources needed for lecanemab or donanemab infusions

